# What Dose of Methamphetamine Do Regular Consumers Use Daily? Estimating Oral Amphetamine Milligram Equivalents

**DOI:** 10.1101/2025.05.09.25327334

**Authors:** Joseph R. Friedman, Adam J. Koncsol, Caitlin A. Molina, Ruby Romero, Jasmine Feng, Michelle Poimboeuf, Morgan E. Godvin, Siddarth Puri, Carla Marienfeld, Chelsea L. Shover

## Abstract

**Introduction:** The purity, accessibility, and affordability of illicit methamphetamine has increased in recent decades, which has been linked to rising rates of methamphetamine-involved overdoses, psychosis, cardiovascular events, and other health consequences. Nevertheless, information about the quantity of methamphetamine used by regular consumers has been limited, despite the potential clinical utility of exposure quantification.

**Methods:** From August 2024-April 2026, self-reported daily methamphetamine consumption was assessed among n=94 individuals in Los Angeles County. Methamphetamine samples (n=256) were analyzed for purity using liquid chromatography-mass spectrometry. Bioavailability by route of administration and stimulant equivalency were obtained from literature. A simulation model leveraging bootstrapping with 1,000,000 draws was used to estimate oral amphetamine milligram equivalents (AME).

**Results:** The average reported methamphetamine consumption was 1.09g daily (95% prediction interval: 0.07-4.00). Average purity was 84.52% (95% PI: 14.90%-100.00%). Given estimated average bioavailability of 66.41% (95% PI: 39.35%-85.24%), the average consumer used an estimated 1,432.54 AME daily (95% PI: 35.74-6,973.05).

**Discussion:** We estimate that consumers of methamphetamine in Los Angeles use an average daily stimulant dose (nearly 1,500 oral AME) about 25-fold higher than the maximum typical recommended clinical dose of mixed amphetamine salts (60mg). This may help explain the limited efficacy of prescription stimulant treatment for methamphetamine use disorder, which typically employs considerably lower quantities. Given this high dose, these findings shed light on the rising incidence of methamphetamine-related sequelae, such as psychosis, cardiovascular complications, and sudden death. Although exposure quantification is commonplace for alcohol and tobacco use disorders, uncertainties in illicit drug markets has complicated this practice for most illicit drugs. This study supports leveraging emerging information from drug checking programs so that clinicians can approximate exposure quantification.

## Introduction

The purity, accessibility, and affordability of illicit methamphetamine has increased in recent decades^1^. This has been linked to rising rates of methamphetamine-involved overdoses, psychosis, psychiatric hospitalizations, cardiovascular events, and other health consequences^2–4^. Nevertheless, information about the actual quantity of methamphetamine used in practice, among people with methamphetamine use disorder, has been limited. This is largely due to the historical difficulty in assessing the purity of illicit substances used by patients. Nevertheless, there is considerable potential clinical utility of exposure quantification, such as understanding how a patient’s methamphetamine use compares to potential replacement doses of prescription stimulants, or assessing the health impacts of dose reductions in illicit stimulant use.

Using recent advancements in drug checking technologies and services, we quantify the purity of methamphetamine samples used by frequent consumers in Los Angeles. We combine this with self-reported daily consumption quantities among the same population to estimate the daily oral amphetamine milligram equivalents (AME). This quantity is likely to be more easily interpretable by clinicians, who are familiar with dosing of mixed amphetamine salts, rather than the quantity of illicit methamphetamine, for which dosing depends on drug concentration, route of administration, and bioavailability.

## Methods

Methamphetamine purity by weight was assessed using liquid chromatography-mass spectrometry (LC-MS) among n=256 samples collected at a community-based drug checking program in Los Angeles County^5^. Samples were limited to those sold as methamphetamine, and for which clients expected only methamphetamine.

Daily methamphetamine consumption volume and route of administration was assessed by self-report using a survey among n=94 individuals who regularly consume methamphetamine (defined as at least last-month use, though most participants used far more frequently) and brought samples for testing.

Bioavailability by route-of-administration and stimulant equivalency factors were drawn from literature review (see supplemental methods).

Daily AME was estimated according to Equation 1, where *Consumption* is the daily milligrams of illicit methamphetamine used, *Purity* is the proportion of active compound in substances sold as illicit “methamphetamine”, *Bioavailability* is the route-of-administration-specific proportion of total drug used that is absorbed systemically, *Methamphetamine: Amphetamine Equivalence* is the ratio of physiological potency between methamphetamine and mixed amphetamine salts, and *Bioavailability Oral Amphetamine Salts* is the fraction of mixed amphetamine salts that is absorbed systemically when taken orally.

*Oral Amphetamine Milligram Equivalents (AME) = Consumption* · *Purity* · *Bioavailability* ·

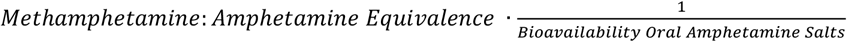

**Equation 1. Calculation of estimated oral amphetamine milligram equivalents (AME) used by regular consumers of methamphetamine in Los Angeles**

To estimate the distribution of AME, combining uncertainty and variation in each of the aforementioned parameter inputs, we employed a bootstrapping approach leveraging 1,000,000 draws. This allows for the inclusion of uncertainty in each parameter, which is propagated through to the final estimated AME. Means and 95% prediction intervals (representing 95% of the estimated values for each parameter) are provided by summarizing across draws.

The UCLA IRB approved this project (IRB-22-0760) and additionally determined that aspects of this work constituted public health surveillance and not human subjects research.

## Results

The average reported daily methamphetamine consumption was 1.09g (95% prediction interval: 0.07-4.00) [Figure 1]. Estimated average purity was 84.52% (95% PI: 14.90%-100.00%).

**Figure 1.**
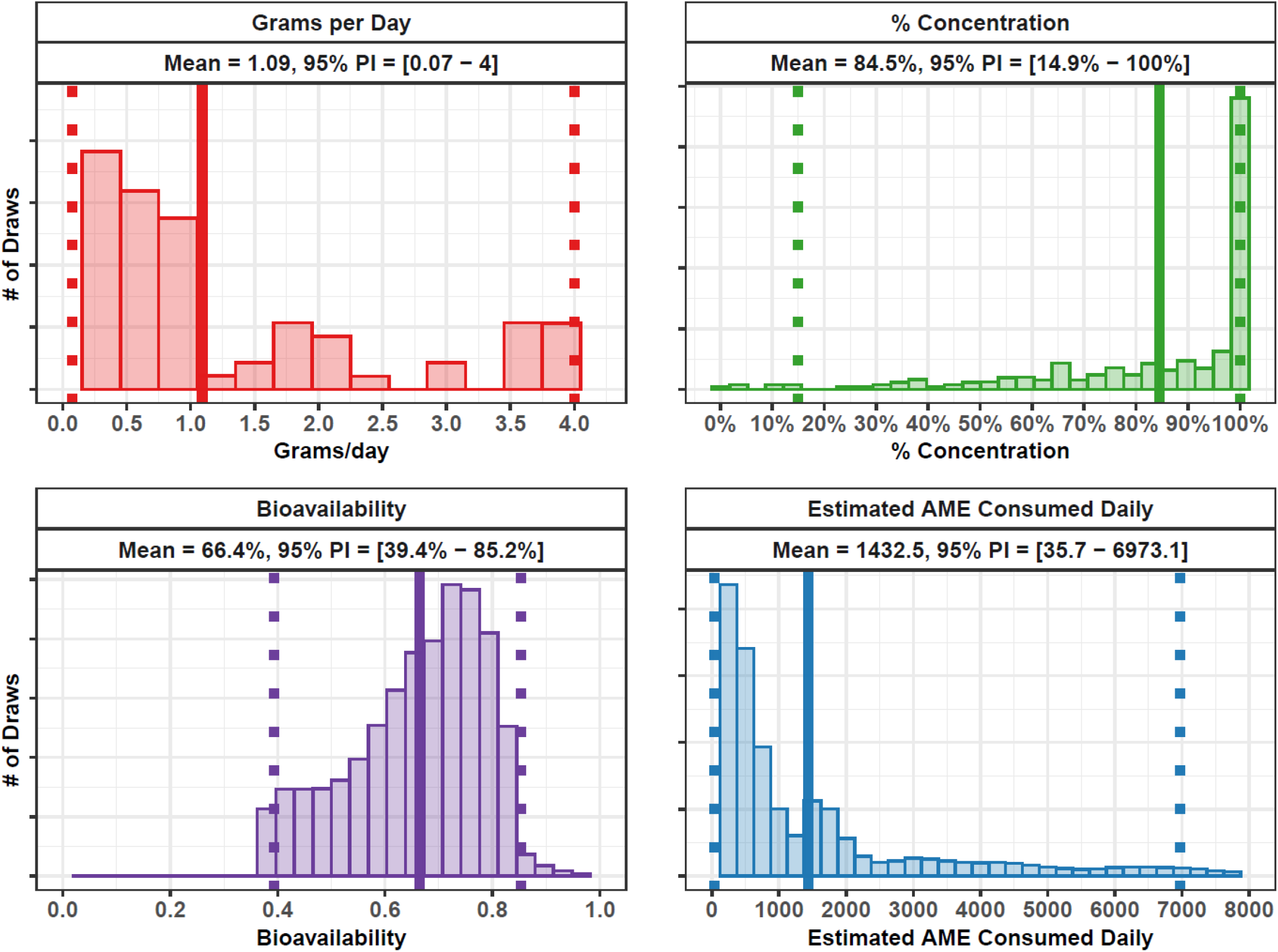
Distributions of Model Parameters: Estimated Daily Methamphetamine Consumption, Percent Concentration (Purity), Bioavailability, and Oral Amphetamine Milligram Equivalents (AME) For each panel, a solid vertical line shows the distribution mean, and a dashed lines shows the prediction interval (representing 2.5^th^ and 97.5^th^ percentiles). Top Left: The distribution of self-reported grams of methamphetamine used per day among regular consumers participating in drug checking services in Los Angeles. Top Right: Percent concentration (purity) of expected methamphetamine samples provided by clients at drug checking services in Los Angeles. Bottom Left: Estimated bioavailability (fraction of drug absorbed into the body) of methamphetamine based on reported routes of administration. Bottom Right: the estimated distribution of oral amphetamine milligram equivalents (AME) consumed among drug checking clients in Los Angeles.

Bioavailability was estimated from literature sources describing experimental data using human subjects (see supplemental methods for a full description). Average bioavailability of smoked methamphetamine was estimated at 52.00% (of the total “pipe dose”), reflecting a uniform distribution with a minimum of 37.00% and a maximum of 67.00%. Bioavailability of insufflated “snorted” methamphetamine was modeled using a normal distribution with a mean of 79.40% and a standard deviation of 6.72%. Oral bioavailability was estimated using a normal distribution with a mean of 67.20% and a standard deviation of 1.59%. Rectal bioavailability was estimated using a normal distribution with a mean of 67.20% and a standard deviation of 20.00%. Bioavailability for injecting was assumed to be 100%. Assuming equal participation in each route of administration when multiple were reported, the average estimated overall bioavailability among study participants was 66.41% (95% PI: 39.35%-85.24%). Based on literature values the amphetamine-methamphetamine equivalency was estimated to be 1:2 (see supplement).

Given the above factors, the average consumer used an estimated 1,432.54 oral AME daily (95% PI: 35.74-6,973.05).

## Discussion

We estimate that consumers of methamphetamine in Los Angeles use an average daily stimulant dose—nearly 1,500 oral amphetamine milligram equivalents (AME)—that is about 25-fold higher than the maximum typical recommended clinical dose of mixed amphetamine salts (60mg). This may help explain the limited efficacy of prescription stimulant treatment for methamphetamine use disorder, which typically employs considerably lower quantities^6^. For instance, studies assessing the efficacy of providing stimulants such as amphetamine salts, as replacement therapy for methamphetamine use disorder, have generally used doses on the order of 60 to 110mg daily^7,8^. Our findings suggest that these doses would represent ∼1/14^th^ to ∼1/25^th^ of the average estimated methamphetamine usage observed in our population.

Given this high dose, these findings also shed light on the rising incidence of methamphetamine-related sequelae, such as psychosis, cardiovascular complications, and sudden death^1,3^, which are much more common with illicit methamphetamine compared to the prescribed use of stimulant therapies^9^.

Although exposure quantification is commonplace for alcohol and tobacco use disorders, uncertainties in illicit drug markets has complicated this practice for most illicit drugs. However, this study suggests that by leveraging emerging information from drug checking programs, as well as regularly asking their patients about consumption quantities, clinicians may be able to approximate exposure quantification. This may be useful in understanding the clinical benefits of treatments that result in dose reductions. It can also inform risk stratification for cardiac, psychiatric and other outcomes and personalizing care, including withdrawal management. This approach would also mirror efforts to quantify use and employ usage reduction as a clinical endpoint in addiction-related clinical trials^10^.

This study represents an example of how emerging drug checking technologies can provide consumers with information that can empower them to make safer drug use decisions, despite the highly uncertain and rapidly evolving nature of the illicit drug supply.

This work is limited by its use of data from a single city and may not be generalizable to other contexts. Drug checking participants represent a convenience sample and may have more severe methamphetamine use disorder than the general population of individuals that use methamphetamine. For instance, in our Los Angeles County sample, smoking was by far the most prevalent route of administration (91.5%), followed by injecting (42.6%) and snorting (36.2%). This high prevalence of smoking aligns with broader regional and national shifts towards this practice^11,12^. However, our cohort demonstrated notably higher rates of injection compared to national averages; e.g. recent national data indicate that only about 13.5% of individuals with lifetime methamphetamine use have injected the drug^13^. The elevated injection rate in our sample (42.6%) likely reflects the nature of our recruitment from a community-based drug checking program based partially at syringe services programs.

Self-report consumption data is also prone to recall and desirability biases. Although the LC-MS testing technology used here is generally considered the ‘gold standard’ for drug content quantification, it can miss certain contaminants not explicitly tested for. LC-MS testing for methamphetamine purity assessment must also be repeated among other parts of Southern California, and other regions, as well as over time, to establish the degree of geographic and temporal variation, which affect the interpretation of these results.

The literature on bioavailability of methamphetamine is also somewhat limited and based on experimental data that is several decades old (see supplemental methods). More updated experiments would provide more precision for this work. Additionally, the comparison between methamphetamine and amphetamine we employ here is imperfect, as methamphetamine has distinct pharmacological properties (methamphetamine’s greater lipid solubility, faster entry into the central nervous system with more rapid onset of action, and higher active d-isomer profile) that affect subjective experience and physical response. The assumption of a 2:1 potency between these two drugs is based on AUC-style exposure estimate—not a pharmacokinetic or reinforcement equivalence and should be treated as a relatively crude measure of equivalence. Nevertheless, given the large magnitude consumption estimated here, bioavailability parameters (or the other potential biases noted above) are unlikely to change our main conclusions.

In sum, we provide the first quantification, to our knowledge, of the daily dose of methamphetamine used by regular consumers. We defined a metric, AME, which can assist clinicians in describing methamphetamine consumption in terms of prescription stimulant equivalent. We find that the average regular consumer uses 25-fold the recommended maximum dose of amphetamine salts, which may explain the myriad and severe health harms observed among patients with long-term methamphetamine use. Further study will be required to assess the degree to which these findings hold true in other populations.

## Data Availability

The complete underlying person-level data leveraged in this study are sensitive and may not be shared. However, summary statistics or other information may be requested from the authors.

## Acknowledgements

The authors report no conflicts of interest. This work was supported by the Centers for Disease Control and Prevention as part of Overdose Data to Action: LOCAL (CDC-RFA-CE-23-0003), and made possible through an equipment grant from the James B. Pendleton Charitable Trust to the UCLA AIDS Institute and UCLA Center for AIDS Research. JRF received funding from the National Institute on Drug Abuse (1U01DA063078). AJK received educational support through the

NIH/National Center for Advancing Translational Science (NCATS) UCLA CTSI (TL1TR001883). CLS received support from the National Institute on Drug Abuse (K01DA050771). The funders played no role in the design and conduct of the study; collection, management, analysis, and interpretation of the data; preparation, review, or approval of the manuscript; and decision to submit the manuscript for publication.

## Supplemental Methods

### Purity

1. Data describing methamphetamine purity and daily quantity ingested among regular consumers were assessed via data generated from anonymous participants accessing a community-based drug checking program, *Drug Checking Los Angeles*. Participants voluntarily provide samples of illicit drug products for testing at four different sites in Los Angeles County, California from August 2024 to April 2026.
2. Samples were analyzed initially in the field with Fourier-transform infrared (FTIR) spectroscopy and immunoassay test strips. Samples were then sent to the National Institute of Standards and Technology (NIST) for secondary laboratory-based qualitative and quantitative testing using direct analysis in real time mass spectrometry (DART-MS) and liquid-chromatography mass spectrometry (LC-MS). Both FTIR and DART-MS assess samples against libraries of over 1,300 substances, including pharmaceutical and illicit drugs, adulterants, cutting and bulking agents, precursor chemicals, and other substances (e.g., adhesives, food products, etc.). The LC-MS quantification panel included twelve substances: fentanyl and fluorofentanyl, fentanyl precursor chemicals (4-ANPP, phenethyl 4-ANPP), heroin, methamphetamine, cocaine, α2-agonists (xylazine and medetomidine), and three common fentanyl adulterants (tetracaine, lidocaine, and Bis(2,2,6,6-tetramethyl-4-piperidyl) sebacate (BTMPS).
3. Of 2,841 total available samples, this study included n=256 that were sold as methamphetamine, expected to contain only methamphetamine, per client self-report, and had quantified results available based on LC-MS.
4. The n=256 purity samples represented at least n=129 unique participants (according to voluntary anonymous identifiers created and provided by participants for future use in accessing their results). There were n=93 samples missing anonymous identifiers because they were collected prior to the implementation of the identifier system or the participant declined to provide one. Of the valid identifiers, one individual provided n=10 samples, two provided n=4, two provided n=3, fifteen provided n=2, and the remaining 109 individuals provided n=1 sample.
5. Methamphetamine purity was initially reported as the freebase form which does not account for the additional weight of the hydrochloride salt form. In order to convert this to methamphetamine HCL values (the form that all samples in our dataset were sold as, and the form that medication forms of stimulants typically consist of) concentration values were multiplied by 1.24 to reflect the ratio of the molecular weights between the HCL and freebase forms of methamphetamine.
6. As a result of measurement limitations, concentration at or above 95% was imputed to be 95%. Additionally, samples with methamphetamine concentration below the limit of quantitation were imputed to be 0.1%.
7. The 256 available methamphetamine purity values were resampled to create 1,000,000 draws representing the distribution of purity using the sample() function in R. A seed was set to ensure the reproducibility of results between code iterations.

### Drug Quantity Consumed

1. Information on drug quantity consumed for methamphetamine (and other drugs) were collected through an anonymous, optional survey conducted by trained drug checking staff.
2. Of n=94 participants who regularly consume methamphetamine (defined as at least past 30 day use) and brought methamphetamine for testing, n= 70 reported the quantity of methamphetamine they consume in grams and had the option to report the quantity per day, week, or month. Weekly values were converted to days by dividing by 7. Monthly values were converted to daily by dividing by 30. N=24 participants reported quantities in dollars spent on methamphetamine per day or week. Dollar values were converted to grams using a standardized price of$20 per gram, which was known to the drug checking team as a typical ‘going rate’ for a gram of methamphetamine. Of note, even cheaper prices (e.g.$10/gram) are often reported by drug checking participants. The used figure of$20 per gram may slightly underestimate the consumption of individuals who spent larger quantities per day and therefore achieve a lower price per gram.
3. The 94 available methamphetamine quantity values (alongside associated route of administration information) were resampled to create 1,000,000 draws representing the distribution of purity using the sample() function in R.

### Bioavailability and Equivalence

1. Literature values were used to estimate credible intervals for all other model parameters, including bioavailability and equivalence factors.
2. Route of administration information was collected alongside consumption quantity information from n=94 participants (see below).

3. Bioavailability of orally consumed methamphetamine was estimated using experimental data from: *Cook, C. E., Jeffcoat, A. R., Sadler, B. M., Hill, J. M., Voyksner, R. D., Pugh, D. E., White, W. R., & Perez-Reyes, M. (1992). Pharmacokinetics of oral methamphetamine and effects of repeated daily dosing in humans. Drug metabolism and disposition: the biological fate of chemicals, 20(6), 856–862* and *Cook CE, Jeffcoat AR, Hill JM, et al. Pharmacokinetics of methamphetamine self-administered to human subjects by smoking S-(+)-methamphetamine hydrochloride. Drug Metab Dispos. 1993;21(4):717-723*. The authors estimated the oral bioavailability of methamphetamine to be 67.20% +/-3.10%. This quantity was used directly as no other studies were found. Draws were estimated using a normal distribution with a mean of 67.20 and a standard deviation of 1.590.
4. Bioavailability of smoked methamphetamine was estimated from three studies:
  a. *Cook CE, Jeffcoat AR, Hill JM, et al. Pharmacokinetics of methamphetamine self-administered to human subjects by smoking S-(+)-methamphetamine hydrochloride. Drug Metab Dispos. 1993;21(4):717-723*. The quantity of interest is the percent of the “total pipe dose” that is ultimately absorbed. This study estimated that a bioavailability of 66% (90% absorption of 73.1% of drug smoked, 26.9% left in pipe).
  b. *Harris DS, Boxenbaum H, Everhart ET, Sequeira G, Mendelson JE, Jones RT. The bioavailability of intranasal and smoked methamphetamine. Clin Pharmacol Ther. 2003;74(5):475-486. doi:10.1016/j.clpt.2003.08.002*. This study estimated a bioavailability of 37.4% (67% absorption of 55% of drug smoked, 45% left in pipe). Nevertheless, the authors used a method that was not designed to maximize the methamphetamine extracted from the pipe, as each participant took only 2 inhalations of each dose. The authors of this study estimate, in their discussion section, that true absorption of an effective pipe dose is likely to be 37% to 67% based on user smoking skill.
  c. *Perez-Reyes, M., White, R., McDonald, S., Hill, J., Jeffcoat, R., & Cook, C. E. (1990). Pharmacologic effects of methamphetamine vapor inhalation (smoking) in man. NIDA research monograph, 105, 575–577*. This study estimated the bioavailability of “vaporized” (i.e., smoked) methamphetamine to be approximately 50%. This was largely a result of analyzing subjective and cardiovascular effects under experimental conditions between IV and smoked methamphetamine. Given the above, the bioavailability of smoked methamphetamine was estimated using a uniform distribution with a minimum of 37.00% and a maximum of 67.00%.
5. The bioavailability of insufflated (“snorted”) methamphetamine was estimated using the following study: *Harris DS, Boxenbaum H, Everhart ET, Sequeira G, Mendelson JE, Jones RT. The bioavailability of intranasal and smoked methamphetamine. Clin Pharmacol Ther. 2003;74(5):475-486. doi:10.1016/j.clpt.2003.08.002*. This study estimated a bioavailability of 79% +/-13.1%. Bioavailability was estimated using a normal distribution with a mean of 79.40 and a standard deviation of 6.72%.
6. No literature sources were found describing the bioavailability of “boofed” or rectally administered methamphetamine. Given that per rectum absorption is often similar or greater than oral absorption, the distribution was estimated using a normal distribution with a mean of 67.2% (the same as for PO bioavailability), yet with a significantly larger standard deviation of 20.00%, given the lack of data on this topic.
7. The bioavailability of injected methamphetamine was, by definition, set at 100%.
8. Among the n=94 individuals providing route of administration information alongside quantity of consumption information (resampled to create 1,000,000 draws) bioavailability was calculated for each draw. If an individual reported multiple routes of administration, then the average of the bioavailability coefficients was used for that draw. In effect, this assumes that individuals evenly split their consumption between the different methods they reported. This could underestimate or overestimate their total absorption depending on their true route of administration habits and the relative frequencies of each method of consumption. If statistical draws for bioavailability distributions exceeded 100%, they were capped at 100%.
9. The methamphetamine to amphetamine potency ratio was assessed using equivalency tables (https://www.adhdmedcalc.com/), which tend to assign a 2:1 ratio, and based on clinical trials data (https://www.accessdata.fda.gov/cdrh_docs/reviews/K040133.pdf) . This value of 2.0 was used for calculations.
10. The bioavailability of oral mixed amphetamine salts was assessed using literature sources. Although no specific experimental data were identified, review sources generally reported a high-level of absorption, with values ranging from 75%-95%. Bioavailability was therefore estimated using a uniform distribution ranging from 75.00% to 95.00%.

| Route of Administration | Number |
| --- | --- |
| Smoke | 29 |
| Smoke, Inject - Iv | 20 |
| Smoke, Snort | 16 |
| Smoke, Inject - Iv, Snort | 8 |
| Snort | 4 |
| Orally Ingest | 2 |
| Smoke, Inject - Iv, Other | 2 |
| Smoke, Inject - Iv, Snort, Orally Ingest, Boof | 2 |
| Smoke, Orally Ingest | 2 |
| Inject - Iv | 1 |
| Inject - Iv, Boof | 1 |
| Smoke, Inject - Iv, Boof | 1 |
| Smoke, Inject - Iv, Inject - Im, Snort, Orally Ingest | 1 |
| Smoke, Inject - Iv, Inject - Im, Snort, Orally Ingest, Boof | 1 |
| Smoke, Inject - Iv, Orally Ingest, Boof | 1 |
| Smoke, Inject - Iv, Snort, Boof, Other | 1 |
| Smoke, Inject - Iv, Snort, Orally Ingest | 1 |
| Smoke, Other | 1 |
| **Total** | **94** |

### Calculations

1. Calculations were performed according to the equation listed in the main text for the 1,000,000 draws of each parameter, derived as defined above. The distribution of quantity consumed, purity, and AME was graphed directly across the draws, and summary statistics were calculated.
2. Means and 95% prediction intervals were calculated for each model parameter. 95% prediction intervals represent the 2.5^th^ and 95.7^th^ percentile of draws, reflecting the bounds within which 95% of the estimated parameter values fall.

